# Citrulline and Faecal Elastase 1 as a Combined Diagnostic Biomarker for Pancreatic Ductal Adenocarcinoma

**DOI:** 10.64898/2026.07.16.26358209

**Authors:** Umar Niazi, Charles AK. Roberts, Declan McDonnell, Victoria M. Goss, Paul R. Afolabi, Jonathan R. Swann, Christopher D. Byrne, Gareth O. Griffiths, Zaed Z. Hamady

**Affiliations:** Cancer Research UK Southampton Clinical Trials Unit, University of Southampton, Southampton SO16 6YD, UK; Department of General Surgery, University Hospital Southampton NHS Foundation Trust, Southampton SO16 6YD, UK; Human Development & Health, University of Southampton, Southampton SO16 6YD, UK; National Institute for Health and Care Research, Biomedical Research Centre, University Hospitals Southampton NHS Foundation Trust, Southampton SO16 6YD, UK

## Abstract

**Background:** Early detection of pancreatic ductal adenocarcinoma (PDAC) is critical. While faecal elastase-1 (FE-1) is a standard clinical marker for pancreatic function, its diagnostic accuracy for malignancy is limited. We sought to identify plasma metabolites that enhance FE-1 performance in symptomatic “at-risk” patients.

**Methods:** Using the DEPEND cohort (CRUK C45617/A29908), plasma metabolomics was performed on patients with resectable PDAC (n=23) and healthy volunteers (n=24). Predictive modelling included feature selection and cross-validation, with further validation in an independent external cohort.

**Results:** Citrulline was identified as significantly depleted in PDAC patients across discovery and validation cohorts. In isolation, Citrulline achieved an AUC of 0.86 (internal) and 0.88 (external validation). Standalone FE-1 demonstrated an AUC of 0.67. However, combining Citrulline and FE-1 significantly improved diagnostic performance, achieving a combined AUC of 0.96. Stratification revealed distinct metabolomic signatures associated with poorly differentiated tumours, suggesting a link to histological grade.

**Conclusions:** Integrating Citrulline with FE-1 testing substantially improves PDAC detection in symptomatic patients. This non-invasive panel offers high diagnostic potential, though prospective validation is required to establish clinical cut-offs for routine practice.

## Introduction

Pancreatic ductal adenocarcinoma (PDAC) remains one of the most lethal malignancies, characterised by an insidious progression and a dismal five-year survival rate of approximately 8% [1]. The median survival for diagnosed patients is frequently less than 12 months [2]. This poor prognosis is primarily driven by the fact that the majority of cases are identified at an advanced stage, where local invasion or distant metastasis precludes potentially curative surgical resection [3]. Conversely, when PDAC is detected at an earlier, resectable stage, the five-year survival rate can increase significantly to over 18% [4]. Consequently, improving long-term outcomes depends heavily on the development of effective strategies for early detection.

Current diagnostic modalities, including imaging and traditional serum protein markers, lack the requisite sensitivity and specificity for population-level or high-risk screening. Carbohydrate Antigen 19-9 (CA 19-9), the most widely utilised biomarker, is frequently elevated in benign hepatobiliary conditions, leading to false positives [5]. Furthermore, approximately 10% of the population are Lewis antigen non-secretors who cannot produce CA 19-9 [6], rendering the marker clinically ineffective for these individuals. Identifying novel, non-invasive biomarkers that can accurately differentiate early-stage PDAC from healthy individuals and benign pancreatic diseases remains a critical unmet clinical need.

Metabolomics, the systematic study of small-molecule metabolites, offers a powerful lens into the metabolic reprogramming that characterises oncogenesis. By analysing shifts in metabolic concentrations, researchers can gain direct insights into the systemic demands and metabolic signatures of tumour progression. Previous studies have documented profound dysregulation in pathways such as glycolysis, the tricarboxylic acid (TCA) cycle, and amino acid metabolism in PDAC [7–9]. These systemic metabolic shifts suggest that small-molecule signatures may circumvent the biological and technical limitations associated with traditional protein-based biomarkers.

The present study utilises data from the Cancer Research UK (CRUK) funded DEPEND trial (CRUK C45617/A29908), a prospective single-centre case-control study conducted at University Hospital Southampton NHS Foundation Trust (UHSFT). The trial was designed to evaluate the diagnostic potential of non-invasive biomarkers in patients presenting with non-specific gastrointestinal symptoms and suspected pancreatic malignancy. The primary cohort involved in this discovery analysis included 47 participants, comprising 23 patients with confirmed resectable PDAC and 24 healthy volunteers. The design and clinical rationale of the study have been detailed previously in the trial protocol [10], while preliminary results regarding the prevalence of pancreatic exocrine insufficiency and the role of faecal elastase-1 (FE-1) in this cohort have been reported elsewhere [11]. Furthermore, the initial metabolomic profiling of this patient group established a baseline for significant metabolite shifts associated with the disease [12].

Adhering to the philosophy that discovery is central to translational research, we leveraged the rich clinical and translational datasets generated by this trial. Central to this effort was the use of the proprietary COSMOS database platform [13]—an integrated SQL schema designed for the management of complex translational data— which enabled the seamless integration of high-dimensional metabolomics profiles with clinical parameters.

In this work, we aimed to transition from the broad characterisation of the PDAC metabolome to the development of a validated, parsimonious predictive signature. Building upon the differential abundance profiles identified in our previous work [12], we employed a rigorous machine learning workflow to identify the most robust individual predictors of the disease. We specifically evaluated the diagnostic potential of Citrulline, and confirmed it using external validation, a metabolite involved in the urea cycle and systemic nitrogen balance and investigated the synergistic effect of combining it with FE-1, an established clinical measure of pancreatic exocrine function. We demonstrate that this combined metabolic-functional signature provides exceptional diagnostic accuracy, offering a promising and clinically translatable pathway toward the early detection of PDAC.

## Methods

### Study Cohort and Sample Collection

This study utilises plasma metabolomics data originally collected as part of the DEPEND trial (CRUK C45617/A29908) [10], a Cancer Research UK-funded prospective case-control pilot study. Participants were recruited from University Hospital Southampton NHS Foundation Trust (UHSFT), who also served as the study sponsor. The cohort comprised male and female patients with histologically confirmed resectable pancreatic ductal adenocarcinoma (PDAC) and a Healthy Volunteer (HV) control group, recruited via a pool of established volunteers via the Project Management Recruitment Team based at UHSFT, or willing friends or family members of the PDAC participants. Characteristics of the DEPEND study participants are presented in Table 1 Ethical oversight was provided by the Health Research Authority and Health and Care Research Wales (REC reference 20/NS/0105, IRAS project ID: 286297).

As the Southampton Clinical Trials Unit (SCTU) is core-funded by Cancer Research UK, this current analysis represents a further exploratory investigation of the collected trial data. This work is specifically aimed at identifying a non-invasive signature for the early diagnosis of PDAC and generating robust hypotheses for future prospective studies. All participants provided written informed consent, and all analyses were undertaken by the SCTU bioinformatics group.

### Metabolomics Analysis

Metabolite abundance was measured in plasma samples using the Biocrates MxP Quant 500 kit (Biocrates Life Science AG, Innsbruck, Austria) at the University of Southampton’s metabolomics facility. This targeted assay allows for the quantification of up to 630 metabolites. The comprehensive analytical procedure, including quality control measures and technical validation, is detailed in our previous characterisation study [11].

### Data Pre-processing and Feature Selection

The discovery dataset comprised metabolite measurements from 47 samples (24 HV and 23 PDAC), supplemented by FE-1 data. To ensure the development of a highly reliable predictive signature, we focused exclusively on metabolites with 100% data completion (i.e., no missing values or concentrations below the limit of detection).

The preliminary differential abundance analysis, comparing metabolite levels between PDAC and HV groups, has been described in detail previously [12]. Briefly, metabolites were filtered based on an adjusted p-value of < 0.05. For the current predictive modelling workflow, we utilised the subset of 136 metabolites that met both the statistical significance threshold and the requirement for complete, non-missing data across all samples. The full list of metabolites and the results of the primary differential abundance analysis are provided in Supplementary Table S1 (Sheet 2). All statistical analyses were conducted using R (version 4.5.2) [14] and the Stan probabilistic programming language (version 2.32.2) [15].

### Predictive Modelling Workflow

To identify a parsimonious biomarker signature and minimize the risk of overfitting in a small sample size, we employed a multi-step feature selection and validation workflow. This approach has been successfully implemented in our previous large-scale “omics” studies to identify robust diagnostic and prognostic signatures [16][17][18]:

#### Validation Set Holdout

A held-out validation dataset of 20 samples (∼43% of the total data) was reserved prior to any training or feature selection steps. This larger-than-standard holdout proportion was chosen to ensure that the validation set contained sufficient statistical power to provide a meaningful estimate of model performance. Furthermore, this allocation was designed to maintain a robust ratio of approximately 10 samples per feature from the minority class, thereby ensuring that the final signature remains stable and less prone to over-fitting on noise within the discovery set.

#### Feature Importance Ranking

We focused on the subset of 136 features previously identified as differentially abundant. We ranked these using two distinct methods:

- **Random Forest:** Using the randomForest package [19] with resampling to estimate mean variable importance.
- **Bayesian GLM with Hierarchical Shrinkage:** A Bernoulli GLM fitted using Stan, employing regularised shrinkage priors to penalize non-predictive coefficients [20].

#### Correlation Pruning

To ensure model parsimony and avoid multi-collinearity, the top-ranked features from both models were merged, and we iteratively removed features with a Pearson correlation coefficient greater than or equal to 0.6.

#### Exhaustive Subset Selection

An exhaustive search was performed on the remaining pruned list to identify the optimal model size (number of predictors) that yielded the smallest test error rate.

#### Performance Estimation

The final selected model was evaluated using Area Under the Curve (AUC) statistics via 10-fold cross-validation on the training set and validated on the reserved holdout data.

### External Validation and Joint Analysis

To validate the generalisability of our findings, we utilised an external dataset reported by Skubisz et al., [21] consisting of serum metabolite profiles from 15 PDAC and 10 healthy control subjects (154 metabolites total). The demographic profile of this validation cohort is detailed alongside our discovery cohort in Table 1. The feature selection workflow previously described—including pairwise correlation reduction and Random Forest/Binomial importance ranking—was applied independently to the Skubisz dataset.

Citrulline, a non-essential amino acid involved in the urea cycle, was identified as a top-ranking feature common to both our internal discovery cohort and the external validation set. To facilitate joint analysis across different laboratory measurement scales, Citrulline values in each dataset were standardised using Z-score scaling. Diagnostic performance was estimated using a binomial model with Citrulline as the sole input. The Area Under the Curve (AUC) was calculated via 10-fold cross-validation on our internal data, followed by an independent validation where the model was trained on our data and tested on the Skubisz cohort.

### Combined Diagnostic Performance: Citrulline and FE-1

We further evaluated the diagnostic utility of Citrulline in combination with FE-1, an established clinical marker of exocrine pancreatic function.

### Clinical Marker Data and Binarisation

A subset of our cohort (N=40; 20 HC and 20 PDAC) had available FE-1 measures. Following clinical guidelines, values <200 µg/g were defined as indicating Exocrine Pancreatic Insufficiency (EPI), while values ≥200 µg/g were considered normal. For model integration, FE-1 was binarised using sum coding (contrast coding): EPI status (<200 µg/g) was assigned a value of -1 and normal status (≥200 µg/g) a value of +1. This centring approach was employed to improve model stability and simplify the interpretation of regression coefficients relative to clinical norms.

### Combined Model Estimation

Diagnostic performance was compared across three models: Citrulline alone, FE-1 alone, and the combined Citrulline/FE-1 signature. All analyses utilised a Bayesian binomial model with a Bernoulli likelihood function implemented in Stan. To address the constraints of a small sample size, we employed shrinkage priors for the regression coefficients to prevent overfitting. Performance was validated using 10-fold cross-validation to estimate the AUC.

### Metabolic Profiling by Tumour Differentiation

To explore metabolic heterogeneity associated with tumour aggression, PDAC patients were stratified by histological grade into ‘Poorly Differentiated’ (N=4) and ‘Moderate-to-Well Differentiated’ (N=9) subgroups.

A Bayesian Generalised Linear Model (GLM) framework was used to estimate average metabolite abundances across these strata; the full results of this subgroup analysis are detailed in Supplementary Table S1 (Sheet 3). To ensure robust estimation, the model utilised a Student’s t-distribution to accommodate heavy tails in the metabolite data. We specifically accounted for unequal within-group variances (heteroscedasticity) by estimating a separate scale term (sigma) at the likelihood level for each group [22]. Cauchy priors (0, 2) were applied to both the regression parameters (betas) and the group-specific scale terms to provide weakly informative regularisation. Critically, metabolites with values below the limit of detection (LOD) were treated as censored data, allowing the model to incorporate information from these observations rather than omitting them. The specific Stan implementations for both the standard and left-censored versions of this heteroscedastic model are provided as source code within Supplementary Table S1 (Sheets 4 and 5).

Due to the limited subgroup sizes, we utilised the model-estimated coefficients (posterior means) to reconstruct robust, “de-noised” average abundance levels. A matrix was constructed using 161 features that were previously identified as significantly differential in the full PDAC vs. HV analysis and were common to the differentiation subgroups.

### Visualisation and Clustering

To visualise patterns of expression across differentiation grades, hierarchical clustering was performed on the reconstructed abundance matrix. Metabolites (rows) were Z-score scaled and clustered using Euclidean distance. The group columns (HC, Poor, and Moderate-to-Well) were clustered using Pearson correlation distance on the unscaled data transpose to preserve the relative magnitude of the group profiles. The resulting heatmap was generated using the NMF R package [23].

## Results

### Differential Abundance and Feature Selection

The differential abundance analysis comparing PDAC and HC groups (HC as baseline), as reported in our previous work, identified 42 under-expressed and 94 over-expressed metabolites, totalling 136 metabolites with a Benjamini-Hochberg adjusted p-value of less than 0.05. Over-expressed metabolites included Di- and Tri-glycerides, Sphingomyelins, Ceramides, Fatty Acids, and Bile Acids. Under-expressed metabolites primarily included Amino Acids and related metabolites. The metabolites with the lowest adjusted p-values included Citrulline, Cortisol, Cer(d18:1/24:1), and 3-Indolepropionic acid.

Pairwise correlation analysis of these 136 metabolites resulted in the removal of 88 features. This high degree of correlation was expected, as many of the removed metabolites were structurally related lipid species (Ceramides, Lysophosphatidylcholines, Phosphatidylcholines, and Triglycerides) sharing common biosynthetic pathways; their removal prevents multi-collinearity issues in downstream modelling.

The remaining 48 metabolites were ranked based on their predictive importance using Random Forest and Binomial GLM. The union of the top 27 ranking candidates from both models included Citrulline, 3-Indolepropionic acid, Cortisol, Glycine, and Butyrylcarnitine among the highest-ranked features. A second round of pairwise correlation analysis removed another 10 metabolites, leaving 17 candidates for exhaustive subset selection.

Exhaustive subset selection indicated that the classification error rate for the test set was lowest for the one-variable model and was closest to the training error rate. As the number of features increased to two and beyond, the training error rate quickly reached zero while the test error rate rose, indicating increased overfitting. Citrulline was identified as the optimal single-candidate biomarker from this exhaustive selection process.

### Reproducibility and Joint Diagnostic Performance of Citrulline

The diagnostic performance of Citrulline, evaluated using ROC analysis and 10-fold cross-validation on our full internal cohort training data, achieved an average AUC of 0.86. To confirm the robustness of this finding, the complete feature selection pipeline was repeated on the external Skubisz et al. dataset [21]. This independent analysis also identified Citrulline among the top-scoring features, alongside others such as Symmetric dimethylarginine, SM (OH) C22:1, and Taurine.

Given the consistent identification of Citrulline across cohorts, showing consistent reduction in Citrulline levels in PDAC patients (Figure 1A), we performed a joint diagnostic analysis using Z-score standardised values. This allowed for a robust performance estimate across different clinical and technological environments. When the model was trained on the full internal cohort and then applied directly to the Skubisz et al. data, it achieved an AUC of 0.88 (Figure 1B). Notably, this cross-cohort AUC replicates the performance reported by Skubisz et al. in their original publication, validating the generalisability of Citrulline as a standalone biomarker.

### FE-1 and Combined Biomarker Performance

Faecal Elastase-1 (FE-1) is a well-established clinical marker of exocrine pancreatic function. In our cohort, 40 subjects provided faecal samples for FE-1 measurement. The diagnostic performance using FE-1 alone (binarised at the 200 ug/g clinical threshold) achieved an average AUC of 0.67 under 10-fold cross-validation (CV).

When Citrulline was analysed alone using the same 10-fold CV on this subset (N=40), it achieved an AUC of 0.86. However, after combining FE-1 and Citrulline into a single predictive model, the 10-fold CV AUC improved significantly to 0.96 (Figure 1C). This result demonstrates a strong synergistic effect between the metabolic marker (Citrulline) and the established clinical marker (FE-1) for PDAC detection.

### Citrulline Distribution and Metabolic Heterogeneity

The distribution of Citrulline in our dataset suggested a larger spread of data in the PDAC group compared to HC. The group-specific scale term (sigma) in our Bayesian GLM was estimated to be approximately two times higher in the PDAC group. This phenomenon—increased variance in the tumour group versus the healthy control—was also observed in the external Skubisz et al. dataset, suggesting it is an intrinsic metabolic characteristic of PDAC and hinting at the presence of distinct metabolic subgroups.

### Metabolic Heterogeneity Driven by Tumour Differentiation

To investigate the source of Citrulline variance, we performed hierarchical clustering of 161 significantly differential metabolites, stratified by histological differentiation grade: Poorly Differentiated (N=4) and Moderate-to-Well Differentiated (N=9). The analysis utilised robust, model-estimated abundance levels derived from the Bayesian GLM accounting for censored data.

The resulting heatmap (Figure 2A) reveals three distinct metabolic phenotypes. While the majority of metabolites show a consistent depletion or enrichment across all PDAC samples, a specific cluster of metabolites exhibits a non-linear expression pattern. This cluster—which includes Citrulline, trans-4-Hydroxyproline (t4-OH-Pro), Arginine, 3- Indolepropionic acid (3-IPA), 1-Methylhistidine, Valine, Homoarginine, Cholesteryl Esters, Creatinine, and alpha-Aminoadipic acid—shows significant suppression in Well-to-Moderately Differentiated tumours compared to HC, but displays a paradoxical “recovery” or elevation in Poorly Differentiated tumours, trending back toward baseline healthy levels (Figure 2B).

## Discussion

In this study, we report a highly effective diagnostic biomarker combination for PDAC, pairing Citrulline (measured via plasma metabolomics) with Faecal Elastase-1 (FE-1), a routine clinical measure of exocrine function. Our feature selection workflow robustly identified Citrulline as the single best-performing metabolite and confirmed its performance in an independent external dataset [21]. Crucially, combining Citrulline with FE-1 significantly enhanced diagnostic performance, achieving an outstanding AUC of 0.96 under cross-validation.

Importantly, Citrulline is not merely a statistical extract from a machine learning algorithm; it is a biologically relevant molecule with a defined role in cancer metabolism. Its selection aligns with a growing body of literature implicating urea cycle dysregulation (UCD) in the pathogenesis and progression of pancreatic cancer [24][25]. This biological grounding provides a critical layer of validation, suggesting that the signal represents a genuine pathophysiological change rather than a cohort-specific artifact.

### Citrulline and the Altered Tumour Microenvironment

Citrulline is a nonessential amino acid, a deiminated form of arginine, and a key intermediate in the Urea Cycle. The three primary enzymes involved in the metabolism of free Citrulline are NO synthase (NOS), ornithine carbamoyl-transferase (OTC), and Argininosuccinate synthetase (ASS1). While the urea cycle is traditionally associated with ammonia detoxification, cancer cells frequently reprogram these pathways to support tumour growth and survival [24]. This UCD phenomenon is thought to promote pyrimidine synthesis by diverting nitrogen toward nucleotide production, providing the essential building blocks for rapid DNA and RNA replication [25].

Specifically, Lee et al. [24] analysed gene expression profiles across 25 cancer types in the TCGA database and found that OTC—the enzyme responsible for producing Citrulline from Ornithine—was consistently downregulated in pancreatic cancer. This downregulation coupled with the observation that poorly differentiated tumours had higher Citrulline levels, suggests that the tumour microenvironment’s attempt to regulate the urea cycle, or perhaps differential enterocyte function due to the tumour burden, is reflected in the blood. Our integration of the Skubisz et al. data confirmed this predictive power (AUC 0.88), further supporting the hypothesis that altered Citrulline levels reflect systemic metabolic reprogramming in PDAC.

### Metabolic Heterogeneity and Tumour Aggression

Beyond global suppression, our sub-analysis revealed a striking, non-linear relationship between the metabolic profile and tumour differentiation. While Citrulline and its co-clustering metabolites—including Arginine, Homoarginine (HArg), and Creatinine—were suppressed in Moderate-to-Well Differentiated tumours, they exhibited a paradoxical “recovery” or elevation in the Poorly Differentiated (aggressive) group.

Although our sample size for this subgroup analysis was small (N=4 vs. N=9), preventing definitive conclusions, this pattern warrants discussion as it mirrors the concept of metabolic plasticity [26]. Aggressive PDAC cells often overcome dependence on external arginine by upregulating enzymes like ASS1 to synthesize their own arginine or recycle Citrulline, supporting rapid proliferation and invasion. The observed elevation of Citrulline, Arginine, and HArg in poorly differentiated tumours may be a systemic signature of this metabolic autonomy.

Furthermore, the co-clustering of trans-4-Hydroxyproline (t4-OH-Pro)—an abundant amino acid in collagen that promotes PDAC invasion [27]—and Creatinine (a marker of muscle and energy metabolism) suggests a combined signature reflecting both tumour aggression and host response. The role of the creatine kinase/phosphocreatine axis is increasingly recognised as an essential energy buffer for rapidly proliferating cells [28]. This suggests that a Citrulline-based signature may not only detect cancer but could offer prognostic insight into tumour grade.

### The Role of Faecal Elastase-1 (FE-1) and the Combined Model

The addition of FE-1 to the model is highly significant because Exocrine Pancreatic Insufficiency (EPI) is extremely common in PDAC due to ductal obstruction and acinar tissue destruction. While FE-1 alone achieved only a modest AUC (0.67), it provides an independent clinical measure of organ dysfunction. The synergistic effect of pairing a metabolic marker (Citrulline) with a functional marker (FE-1) is the key to achieving our high-accuracy results (AUC 0.96).

### Comparison with Existing Biomarkers

This combination offers a significant advantage over the established marker, CA 19-9. While CA 19-9 is widely used, its diagnostic value is modest, with meta-analyses reporting an average AUC of 0.84 [29]. Furthermore, CA 19-9 suffers from low specificity in benign biliary conditions and is unreliable in the 10% of the population who are Lewis-negative. In contrast, our model is based on fundamental metabolic and functional changes. Future prospective studies should compare this two-marker panel head-to-head with CA 19-9.

### Limitations and Future Directions

Despite the promising results, our study has several limitations. First, the sample size (N=47) is modest, and the findings are from a single-centre cohort. Second, the retrospective nature means the links are correlative rather than causal. Third, we did not perform functional in vitro studies to fully elucidate the mechanism of Citrulline alteration.

Future research should focus on:

1. **Prospective Validation:** Conducting a large-scale, multi-centre prospective cohort study to validate the AUC 0.96 in a general population, potentially focusing on high-risk individuals (e.g., new-onset diabetes).
2. **Mechanistic Studies:** Investigating the underlying mechanism through cell line and animal models, specifically examining the regulation of urea cycle enzymes (OTC, ASS) by PDAC cells.
3. **Early Detection:** Exploring the diagnostic value of this panel in patients with precursor lesions (e.g., IPMNs) to determine its utility as a true early-detection screening tool.

### Clinical Implications

The diagnostic performance of the Citrulline/FE-1 panel suggests a promising, translatable clinical tool. Following further evaluation, this non-invasive panel could be implemented to stratify high-risk patients or evaluate symptomatic individuals, enabling earlier diagnosis when more treatment options exist, thereby impacting the dismal prognosis of PDAC.

## Acknowledgements

The authors would like to acknowledge the hepatopancreaticobiliary (HPB) surgeons and extended members of the HPB team at UHSFT for their assistance in recruiting participants for this trial.

## Authors’ contributions

- **Umar Niazi**: Planning and conducting the study, bioinformatics and statistical analysis, and drafting the manuscript.
- **Charles AK. Roberts**: Data collection and analysis, interpretation of results, and drafting the manuscript.
- **Declan McDonnell, Victoria M. Goss, Paul R. Afolabi, and Jonathan R. Swann**: Data collection, technical support, and critical revision of the manuscript.
- **Christopher D. Byrne, Gareth O. Griffiths, and Zaed Z. Hamady**: Conceptualisation, study design, supervision, and critical revision of the manuscript for important intellectual content.
- Statement: All authors have approved the final draft submitted.

## Ethics approval and consent to participate

The study was conducted in accordance with the Declaration of Helsinki and approved by the Ethics Committee of the Health Research Authority (HRA) and Health and Care Research Wales (HCRW) to be conducted in (and sponsored by) the University Hospital Southampton NHS Foundation Trust (UHSFT) (IRAS project ID: 286297, REC reference 20/NS/0105, on 21 October 2020). Informed consent was obtained from all subjects involved in the study.

## Consent for publication

This study used fully anonymised data, and no identifiable individual level information presented in the manuscript. All data were anonymised prior to analysis by the and no identifiable information was accessible to the research team.

## Data availability

The data that support the findings of this study are available from the corresponding author, [U.N.], upon reasonable request. Supporting data for external validation can be downloaded at https://www.mdpi.com/article/10.3390/cancers15123242/s1.

## Guarantor of the article

Zaed Z. Hamady and Gareth O. Griffiths accept full responsibility for the conduct of the study. They had access to the data and had control of the decision to publish.

---

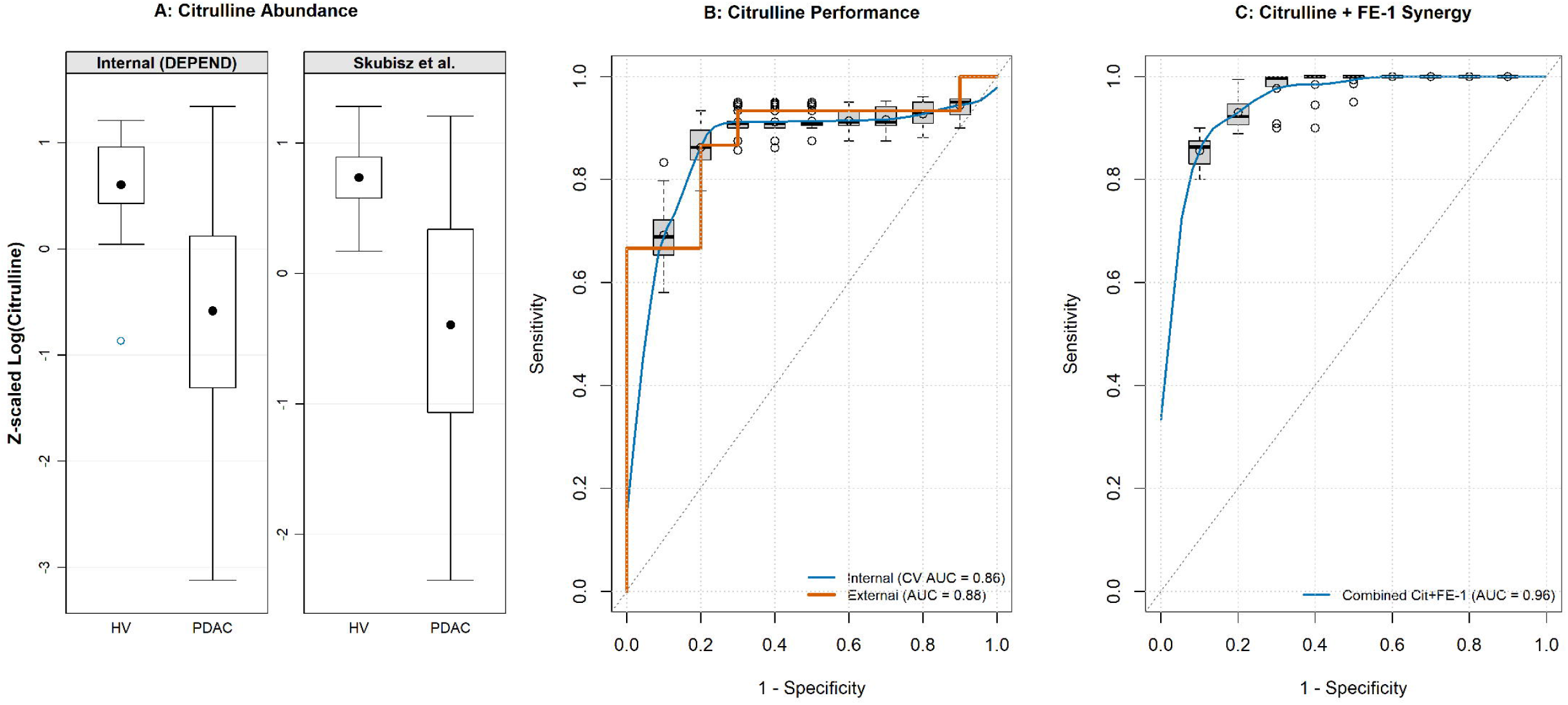

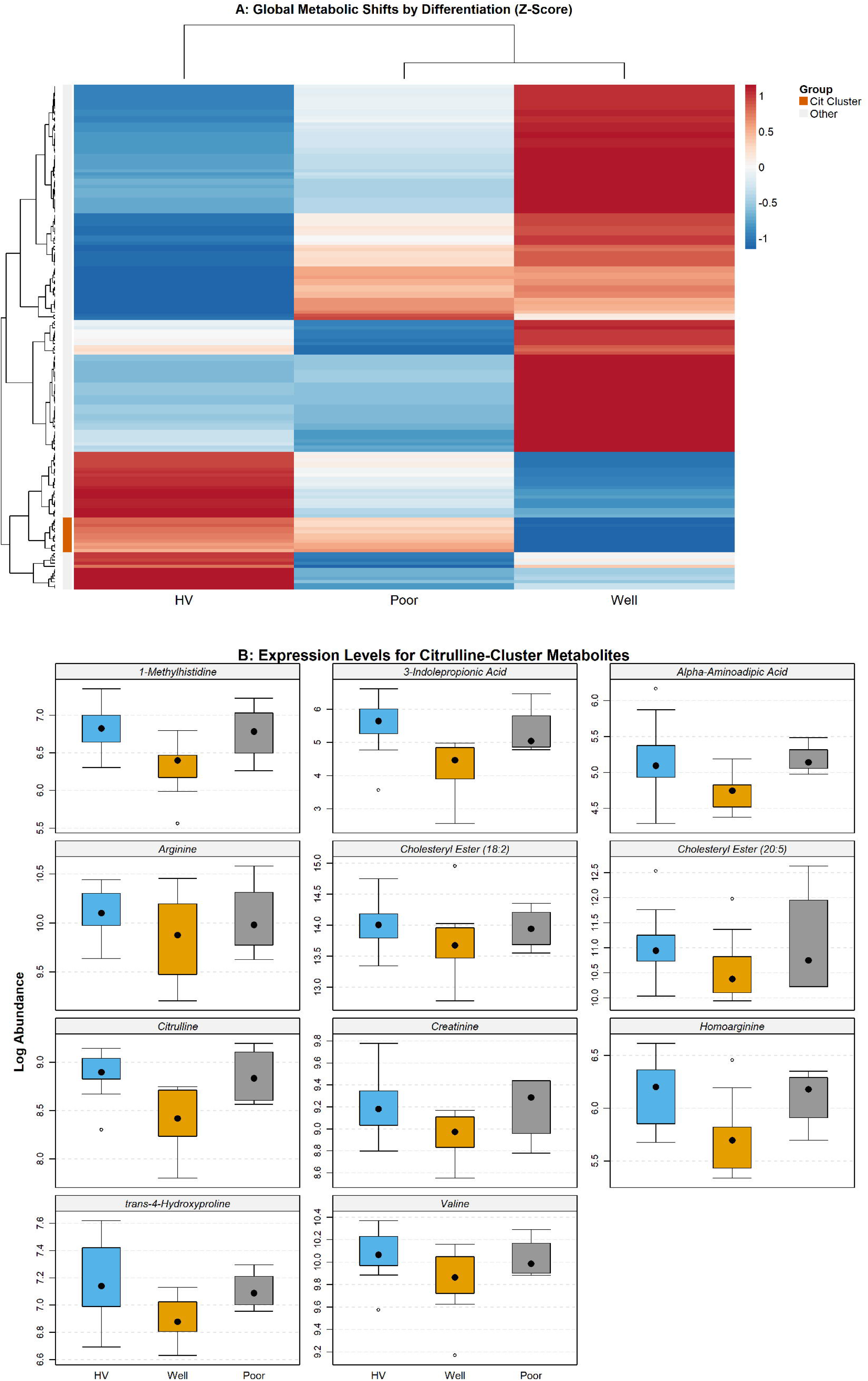

