## Supplementary material for "Citrulline and Faecal Elastase 1 as a Combined Diagnostic Biomarker for Pancreatic Ductal Adenocarcinoma": Tables

**Table 1:** Clinical and demographic characteristics of the study participants. The table compares Healthy Volunteers (HV) and Pancreatic Ductal Adenocarcinoma (PDAC) patients from the internal DEPEND study, alongside the external Skubisz et al. validation cohort.

| **Participant Characteristics** | **DEPEND Healthy Volunteers**  **(n=24)** | **DEPEND** **PDAC**  **(n=23)** | ***P-*value** | **Skubisz et al**  **PDAC**  **(n=15)** |
| --- | --- | --- | --- | --- |
| **Age (Years)** | 62.5 (58.0 - 71.0) | 68.0 (59.5 - 74.5) | 0.495 | 66 (58.0–75.5) |
| **Sex (Male)** | 13 (54.2%) | 16 (69.6%) | 0.371 | 8 (53.3%) |
| **Weight (kg)** | 79.7 (64.6 - 91.6) | 79.0 (71.3 - 83.5) | 0.647 | 62 (50.0 - 71.5) |
| **Height (cm)** | 168.5 (163.0 - 174.1) | 173.5 (168.5 - 179.0) | 0.139 | 165.0 (158.0–171.0) |

Note: Continuous variables (Age, Weight, Height) are presented as Median (Interquartile Range). Categorical variables (Sex) are presented as n (%).

*P*-values represent the comparison between the DEPEND HV and DEPEND PDAC groups, calculated using the Wilcoxon Rank Sum test for continuous data and Fisher’s Exact test for categorical data. A *p-*value < 0.05 was considered statistically significant.
