## Supplementary material for "Citrulline and Faecal Elastase 1 as a Combined Diagnostic Biomarker for Pancreatic Ductal Adenocarcinoma": Figure Legends

**Figure 1. Performance of Citrulline as a Standalone and Synergistic Biomarker.**

**(A)** Distribution of Z-score standardized Citrulline levels across the internal discovery cohort (DEPEND) and the external validation cohort (Skubisz et al.). PDAC patients consistently show lower levels of Citrulline compared to healthy volunteers (HV) across both cohorts. **(B)** ROC analysis of Citrulline performance. The model trained on internal data (Blue; AUC = 0.86) generalizes effectively when applied to the external Skubisz dataset (Orange; AUC = 0.88). **(C)** Synergistic diagnostic performance of the combined model. Integrating plasma Citrulline with binarized Faecal Elastase-1 (FE-1) measures significantly improves diagnostic accuracy (AUC = 0.96) compared to either marker alone.

**Figure 2. Metabolic Heterogeneity Driven by Histological Differentiation.**

**(A)** Heatmap representing the posterior mean abundance of 161 significantly differential metabolites, stratified by Healthy Controls (HV) and PDAC histological grade (Moderate-to-Well vs. Poorly Differentiated). Data are Z-score scaled by row; blue indicates lower abundance and red indicates higher abundance. The 'Citrulline-Cluster' is highlighted in orange. **(B)** Log abundance profiles of the "Citrulline-Cluster" metabolites identified via hierarchical clustering. These metabolites (including Citrulline, Arginine, and 3-Indolepropionic acid) show significant suppression in moderate-to-well differentiated tumors but exhibit a paradoxical recovery toward baseline levels in poorly differentiated (aggressive) PDAC, suggesting a shift in metabolic autonomy as tumor grade increases.
